# Understanding the pathways leading to socioeconomic inequalities in HIV testing uptake in 18 sub-Saharan African countries: a mediation analysis

**DOI:** 10.1101/2021.09.18.21263768

**Authors:** Pearl Anne Ante-Testard, Mohamed Hamidouche, Bénédicte Apouey, Rachel Baggaley, Joseph Larmarange, Tarik Benmarhnia, Laura Temime, Kévin Jean

**Author notes:** Corresponding author: Pearl Anne Ante-Testard, Laboratoire MESuRS, Conservatoire national des Arts et Métiers, 292 rue Saint Martin, 75003, Paris, France.

## Abstract

**Introduction:** Although socioeconomic inequalities in HIV prevention, testing and treatment services have been well documented, their drivers remain poorly understood. Understanding the different pathways between socioeconomic position and HIV testing across different countries could help designing tailored programs aimed at reducing such inequalities.

**Methods:** We analysed data from Demographic and Health Surveys conducted between 2010 and 2018 in 18 sub-Saharan African countries (Burkina Faso, Cameroon, Côte d’Ivoire, Congo DR, Ethiopia, Guinea, Kenya, Lesotho, Liberia, Malawi, Mali, Niger, Rwanda, Senegal, Sierra Leone, Tanzania, Zambia and Zimbabwe). Using a potential outcomes framework and the product method, we decomposed the total effect linking wealth and recent (< 12 months) HIV testing into i) direct effects, and ii) indirect effects, via *demand-related* (related to individual’s ability to perceive need for care and inclination to seek care) or *supply-related* (related to individual’s ability to reach, pay for and engage in health care) mediators. Multivariable gender-specific modified Poisson models were fitted to estimate proportions mediated, while accounting for exposure-mediator interaction when present.

**Results:** A total of 392,044 participants were included in the analysis. Pro-rich wealth-related inequalities were observed in a majority of countries, with nine countries with high levels of inequalities among women and 15 countries among men.

The indirect effects of each mediator varied greatly across countries. The proportion mediated tended to be higher for *demand-related* than for *supply-related* mediators. For instance, among women, HIV-related knowledge was estimated to mediate up to 12.1% of inequalities in Côte d’Ivoire; this proportion was up to 31.5% for positive attitudes toward people living with HIV (PLHIV) in Senegal. For the four *supply-related* mediators, the proportion mediated was systematically below 7%. Similar conclusions were found when repeating analyses on men for the *demand-related* mediators, with higher proportions mediated by positive attitudes toward PLHIV (up to 39.9% in Senegal).

**Conclusions:** Our findings suggest that socioeconomic inequalities in HIV testing may be mediated by the *demand-side* more than *supply-side* characteristics, with important variability across countries. Overall, the important inter-country heterogeneity in pathways of socioeconomic inequalities in HIV testing illustrates that addressing inequalities requires tailored efforts as well as upstream interventions.

A French version of the abstract is available upon request from the corresponding author.

## Introduction

HIV continues to affect many lives globally especially in sub-Saharan Africa (SSA) which accounts for 59% of new HIV infections in 2019 [1] making HIV prevention and treatment essential, particularly in this region. HIV testing has played a crucial role in the prevention and management of HIV/AIDS as the entry point that links individuals to prevention and treatment services.

Routine offer of HIV testing in health settings, such as antenatal clinics was recommended by the World Health Organization in 2007 [2], which changed the profiles of testing users and increased uptake in HIV testing [3]. However, in spite of the significant progress in reducing HIV incidence over the past decade in SSA, HIV incidence has not declined sufficiently to reach the UNAIDS 90-90-90 fast-track goals by 2020 and the Sustainable Development Goal of ending the AIDS epidemic by 2030 [4]. A modelling study that investigated the progress towards the first 90 (i.e., 90% of PLHIV will know their status) found that 84% of PLHIV in SSA knew their status by 2020, with proportions consistently lower in Western and Central Africa (WCA, 67% and 70%, respectively) than in Eastern and Southern Africa (ESA, 86% and 90%, respectively) [5]. These left a gap of around 3.8 million PLHIV left undiagnosed in SSA [5].

Health inequalities that favour the wealthiest subgroups have also persisted in most SSA countries, especially in WCA [6]. Studies found that people with higher socioeconomic position (SEP), and those who were employed, living in urban areas and had heard about HIV and AIDS were associated with better knowledge of HIV status and were more likely to seek testing [7–13]. Potential drivers of these inequalities include HIV-related knowledge, HIV stigma, distance to care and cost of services, among many others. A study found that cost of services, and physical distance between health facilities and service user’s residence were the most significant supply-side barriers in accessing obstetric care in sub-Saharan Africa [14]. We hypothesize these drivers to also be important barriers in accessing HIV testing services. Documenting such mechanisms can be useful in understanding the role of each factor in driving such inequalities.

Despite the literature in socioeconomic inequalities in HIV testing, few studies have explored their possible underlying mechanisms. Such studies are timely though to help better orientate testing strategies in order to reach the first 95 of the 2030 UNAIDS 95-95-95 targets and to ensure “no one is left behind”. In this study, we analysed population-based surveys to understand mediating factors linking SEP and HIV testing uptake at the individual level.

## Methods

### Data and Study Design

We analysed data from the Demographic and Health Surveys (DHS) conducted between 2010 and 2018 to understand the role of different mediating factors in the pathway between SEP and recent (< 12 months) HIV testing uptake.

The DHS are publicly available nationally representative population-based surveys, conducted regularly in low- and middle-income countries (LMIC), collecting data on a wide range of objective and self-reported health indicators including data on HIV/AIDS, using a two-stage sampling design [15]. All women aged 15-49 years are all eligible in all households and, in some surveys, men aged 15-54/59 from a sub-sample are also eligible to participate (https://dhsprogram.com/). The national implementing agencies or research institutes that conducted the surveys were responsible for ethical clearance which assured informed consent from the participants prior to their involvement and guaranteed confidentiality of information [16]. Those who consented are interviewed face-to-face by trained interviewers using a standardized questionnaire that includes items on different sociodemographic characteristics, maternal and reproductive health, and HIV-related questions [15].

Country sample was based on convenience sampling (data available as of February 2021) that was slightly extended from a previous study [6]. In total, we analysed 10 WCA countries (Burkina Faso, Cameroon, Côte d’Ivoire, Cameroon, Congo DR, Guinea, Liberia, Mali, Niger, Senegal and Sierra Leone) and eight ESA countries (Ethiopia, Kenya, Lesotho, Malawi, Rwanda, Tanzania, Zimbabwe and Zambia).

### Variables

#### Socioeconomic Position

We defined participant SEP based on the DHS wealth index, a composite measure of household wealth based on living standards such as household assets and characteristics [17]. More specifically, we used the wealth rank of the participants in the country-specific cumulative distribution of the wealth index, a continuous variable ranging from 0 to 1.

#### Outcome variable

The outcome of interest was the self-report of recent (< 12 months) HIV testing.

#### Mediators

We selected six potential mediators available in the DHS that we hypothesized to be in the pathway between wealth and recent HIV testing uptake based on the literature. We categorized these mediators in two categories based on a principal component analysis for women (Figure S1). The first category of factors referred to the individual’s ability to perceive the need for care and inclination to seek care [18] (i.e., HIV-related knowledge and positive attitudes toward PLHIV). The second mediators’ category included factors that characterize the ability to reach, pay for, and engage in health care [18] (i.e., reporting no distance-related problem to seek care, reporting no money-related problem to seek care, no permission needed from spouse/partner to seek a doctor and no/single difficulty in seeking care). For simplicity, we labelled the first category *demand-related* and the second category *supply-related* mediators. *Supply-related* variables were only available for women in the DHS.

All mediators were coded as binary variables with favourable responses coded as 1. Complete descriptions of these variables and how they were constructed can be found in Table S1.

#### Confounders

The confounders that we identified a priori were age (15-24, 25-34, 35 and above), type of residence (urban and rural) and family situation (in a union, single and widowed/separated).

### Statistical analysis

For each country, we estimated the proportion of the favourable level of each mediator among the richest and the poorest wealth quintiles while accounting for the survey design.

To compute the effect estimates, we fitted multivariable modified Poisson regressions while accounting for the survey design and adjusting for confounders [19]. We estimated socioeconomic inequalities in recent HIV testing uptake by estimating the total effect (TE) of wealth on recent testing (Equation 1).

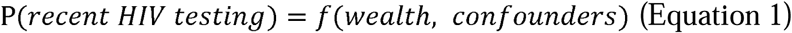

Outcome model:

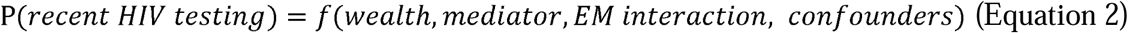

Mediator model:

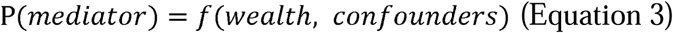

We applied different mediator and outcome models (Equations 2 and 3) using the product method based on the potential outcomes framework [20,21] to explore different pathways linking wealth and recent HIV testing uptake through *demand-related* and *supply-related* mediators. We assumed that these mediators of interest to do not influence one another in the analysis. We considered four assumptions in this analysis: (1) no unmeasured exposure – outcome confounding, (2) no unmeasured mediator – outcome confounding, (3) no unmeasured exposure – mediator confounding, and (4) none of the mediator – outcome confounder is itself affected by the exposure [22]. Figure 1 shows the Directed Acyclic Graph (DAG) of the pathway and effect estimates that we are interested in.

**Figure 1.**
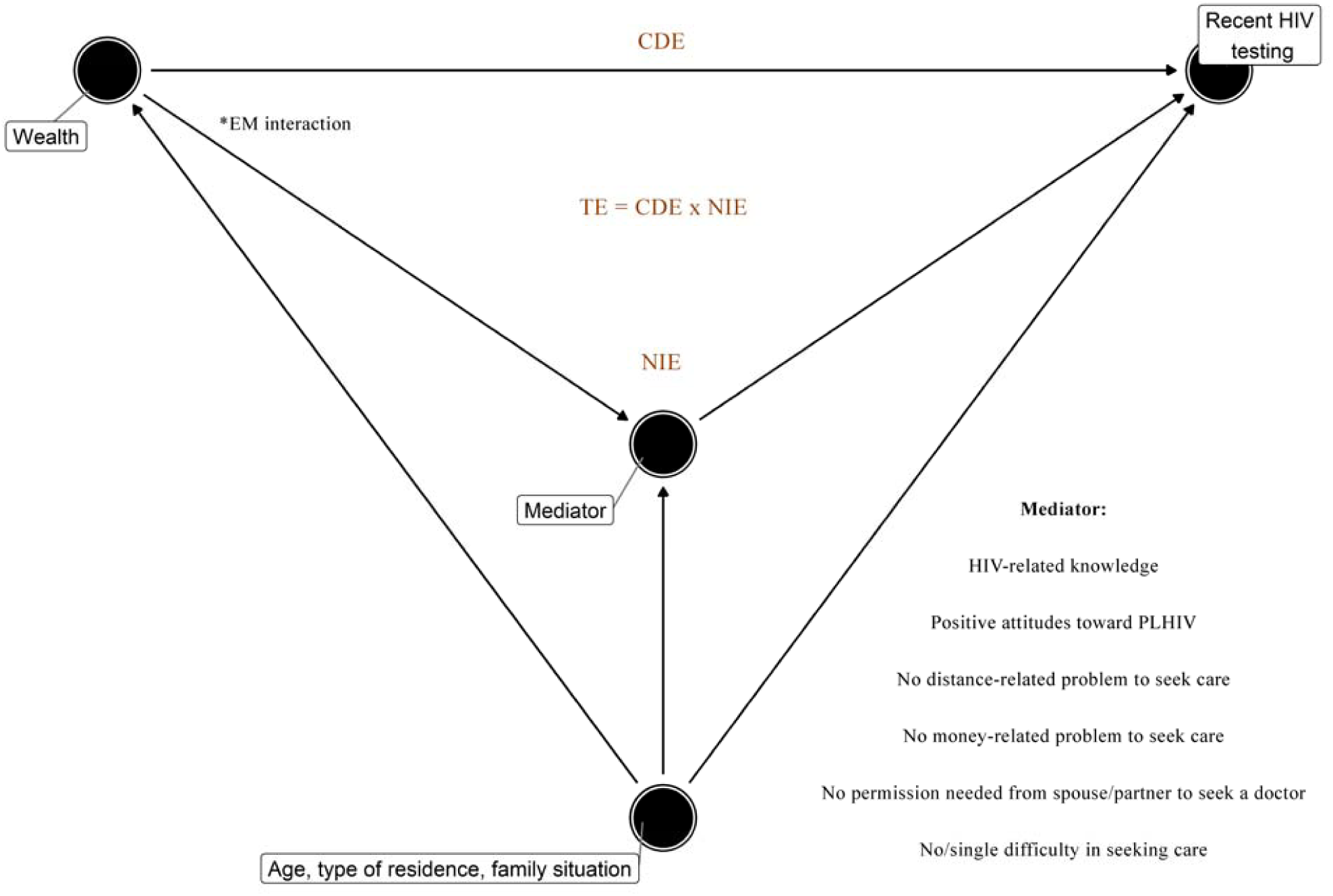
DAG illustrating the pathway between wealth and recent (< 12 months) HIV testing through each mediator while adjusting for confounders (i.e., age, type of residence and family situation) and exposure-mediator interaction when present (*). The TE of wealth on recent HIV testing could be disentangled into the CDE and the NIE. TE: Total Effect. CDE: Controlled Direct Effect. NIE: Natural Indirect Effect. EM: Exposure-mediator.

The TE of wealth on recent HIV testing was decomposed into the controlled direct effect (CDE) and the natural indirect effect (NIE). By using coefficients from the outcome and mediator models (Equations 2 and 3, respectively), we estimated the CDE and NIE. The CDE is the effect of the exposure on the outcome, while the mediator is set to a pre-specified level uniformly over the entire population [21]. Here, we pre-specified the level of the mediator to a favourable level. The NIE represents the change in the outcome when SEP is held constant (X=x) and the mediator changes to what it would have been for a change in the other SEP category (X=x*) [21,23].

We also accounted for exposure-mediator interaction when present to calculate the CDE and NIE by following the formulas from Valeri and Vanderweele [21] (Text S1). By using the CDE and NIE, we estimated the proportion mediated (PM, in %) which is the proportion of the total effect of the exposure on the outcome that is mediated (Text S1). The PM captures how important the pathway is through the mediator in explaining the observed effect of the exposure on the outcome (i.e., TE) [24].

In order to focus on settings in which socioeconomic inequalities were substantial before decomposing such effects into CDE and NIE, we established a cut-off based on the total effect, with a Prevalence Ratio (PR) ≥ 1.5.

We bootstrapped the 95% confidence interval (CI) with 1000 replications. All analyses were conducted using R version 4.0.3.

## Results

### Characteristics of Study Population

Data were collected from 392,044 participants, 261,935 female participants and 130,109 male participants. Table 1 and Table S2 illustrate the survey and participants characteristics while accounting for survey design and sampling weights. Overall, 93-100% of eligible women were successfully interviewed, and 86-100% of men (Table S2).

**Table 1.** Survey and population characteristics, by country and gender.

| Western-Central Africa | BFA<br>(Burkina Faso) |  | CIV<br>(Côte d'Ivoire) |  | CMR<br>(Cameroon) |  | COD<br>(Congo DR) |  | GNA<br>(Guinea) |  | LIB<br>(Liberia) |  | MLI<br>(Mali) |  | NIG<br>(Niger) |  | SEN<br>(Senegal) |  | SLE<br>(Sierra Leone) |  |
| --- | --- | --- | --- | --- | --- | --- | --- | --- | --- | --- | --- | --- | --- | --- | --- | --- | --- | --- | --- | --- |
|  | 2010 |  | 2011-12 |  | 2018 |  | 2013-14 |  | 2018 |  | 2013 |  | 2018 |  | 2012 |  | 2017 |  | 2013 |  |
|  | F | M | F | M | F | M | F | M | F | M | F | M | F | M | F | M | F | M | F | M |
| N | 17,087 | 7,307 | 10,060 | 5,135 | 14,677 | 6,978 | 18,827 | 8,656 | 10,874 | 4,117 | 9,239 | 4,118 | 10,519 | 4,618 | 11,160 | 3,928 | 16,787 | 6,977 | 16,658 | 7,262 |
| HIV-related knowledge (%) | 23.3 | 21.5 | 14.5 | 13.2 | 32.4 | 25.8 | 11.8 | 13.8 | 14.6 | 18.9 | 1.3 | 17.1 | 15.8 | 16.3 | 10.0 | 13.9 | 16.2 | 21.2 | 21.9 | 14.0 |
| Positive attitudes toward PLHIV (%) | 32.8 | 38.7 | 47.3 | 46.3 | 57.0 | 51.2 | 33.6 | 40.2 | 15.4 | 17.5 | 33.6 | 37.5 | 29.5 | 34.0 | 18.3 | 24.9 | 34.2 | 30.0 | 34.7 | 33.0 |
| No distance-related problem to seek care (%) | 56.4 | - | 60.3 | - | 60.3 | - | 61.1 | - | 53.9 | - | 59.9 | - | 71.5 | - | 57.1 | - | 77.9 | - | 61.4 | - |
| No money-related problem to seek care (%) | 28.2 | - | 33.0 | - | 32.7 | - | 31.4 | - | 39.9 | - | 53.1 | - | 59.5 | - | 40.1 | - | 55.3 | - | 32.9 | - |
| No permission needed to see a doctor (%) | 78.9 | - | 75.6 | - | 65.4 | - | 67.3 | - | 70.5 | - | 92.2 | - | 72.9 | - | 78.9 | - | 93.4 | - | 82.5 | - |
| No/ single difficulty in seeking care (%) | 56.6 | - | 56.7 | - | 53.5 | - | 54.1 | - | 54.2 | - | 70.6 | - | 68.1 | - | 59.2 | - | 80.4 | - | 59.7 | - |
| Recent (<12 months) HIV testing (%) | 11.8 | 8.6 | 15.4 | 9.9 | 40.0 | 35.0 | 9.1 | 7.6 | 9.4 | 5.9 | 21.6 | 13.7 | 9.2 | 4.9 | 8.4 | 2.7 | 13.0 | 6.3 | 17.6 | 8.2 |
| Eastern-Southern Africa | ETH<br>(Ethiopia) |  | KEN<br>(Kenya) |  | LES<br>(Lesotho) |  | MWI<br>(Malawi) |  | RWA<br>(Rwanda) |  | TNZ<br>(Tanzania) |  | ZBW<br>(Zimbabwe) |  | ZMB<br>(Zambia) |  |  |  |  |  |
|  | 2016 |  | 2014 |  | 2014 |  | 2015-16 |  | 2014-15 |  | 2011-12 |  | 2015 |  | 2018 |  |  |  |  |  |
|  | F | M | F † | M | F | M | F | M | F | M | F | M | F | M | F | M |  |  |  |  |
| N | 15,683 | 12,688 | 31,079 | 12,819 | 6,621 | 2,931 | 24,562 | 7,478 | 13,497 | 6,217 | 10,967 | 8,352 | 9,955 | 8,396 | 13,683 | 12,132 |  |  |  |  |
| HIV-related knowledge (%) | 18.3 | 27.0 | 28.6 | 23.7 | 27.0 | 16.5 | 35.6 | 31.5 | 44.8 | 40.3 | 30.5 | 25.5 | 44.6 | 34.9 | 33.7 | 24.9 |  |  |  |  |
| Positive attitudes toward PLHIV (%) | 35.5 | 44.5 | 73.0 | 77.7 | 84.4 | 71.6 | 80.5 | 85.0 | 83.4 | 86.4 | 59.1 | 64.8 | 77.2 | 78.8 | 70.7 | 75.1 |  |  |  |  |
| No distance-related problem to seek care (%) | 49.7 | - | 77.3 | - | 74.5 | - | 44.4 | - | 78.4 | - | - | - | 66.7 | - | 28.8 | - |  |  |  |  |
| No money-related problem to seek care (%) | 45.2 | - | 63.3 | - | 72.7 | - | 47.2 | - | 50.7 | - | - | - | 57.0 | - | 20.5 | - |  |  |  |  |
| No permission needed to seek care (%) | 67.9 | - | 94.0 | - | 96.4 | - | 83.6 | - | 97.3 | - | - | - | 94.7 | - | 3.8 | - |  |  |  |  |
| Single/no difficulty in seeking care (%) | 55.1 | - | 82.0 | - | 84.9 | - | 56.2 | - | 82.8 | - | - | - | 76.4 | - | 85.2 | - |  |  |  |  |
| Recent (<12 months) HIV testing (%) | 21.2 | 19.7 | 67.9 | 57.5 | 59.1 | 37.9 | 44.4 | 42.3 | 39.7 | 36.9 | 32.5 | 27.7 | 49.3 | 36.8 | 65.4 | 53.4 |  |  |  |  |

Abbreviations: F, female; M, male, N, total number.
† Women: 31,079 (15-49 years of age: all version); Women: 14,741 (15-49 years of age: full version); Women: 16,338 (15-49 years of age: short version).

In many of the countries, female and male participants lived in rural areas (except in Côte d’Ivoire, Cameroon and Liberia among both genders, and in Senegal among males). They were either married or cohabitating except in Cameroon, Senegal and Lesotho where most males were single (Table S2).

Table 1 shows that around 18%-45% of the female participants and 17%-40% of the male participants had comprehensive HIV-related knowledge in ESA countries compared to 1%-32% and 13%-26% among female and male participants, respectively, in WCA countries. Moreover, the proportion of participants with positive attitudes toward PLHIV were lower in WCA countries (around 15%-57% among females and 18%-51% males) compared to ESA countries (about 36%-84% among females and 45%-86% among males). In terms of the *supply-related* variables, most women reported no *supply-related* problems except in Burkina Faso, Côte d’Ivoire, Cameroon, Congo DR, Guinea, Niger and Sierra Leone in WCA (where a majority of women reported money-related problems in seeking care), and in Ethiopia, Malawi and Zambia in ESA (where a majority of women reported distance-related and money-related problems in seeking care). Most female participants did not need permission from spouse/partner to seek a doctor in all countries except in Zambia.

Self-reported recent HIV testing uptake among female and male participants in WCA was lowest in Niger (8.4% and 2.7%, respectively) and highest in Cameroon (40% and 35%, respectively). Meanwhile in ESA, uptake among women and men was lowest in Ethiopia (21.2% and 19.7%, respectively) and highest in Kenya (67.9% and 57.5%, respectively).

### Socioeconomic Inequalities in HIV Testing

Figure 2 illustrates that the richest were more likely to have comprehensive HIV-related knowledge, have positive attitudes toward PLHIV and were less likely to report *supply-related* problems. We also observed different magnitudes across countries and mediators.

**Figure 2.**
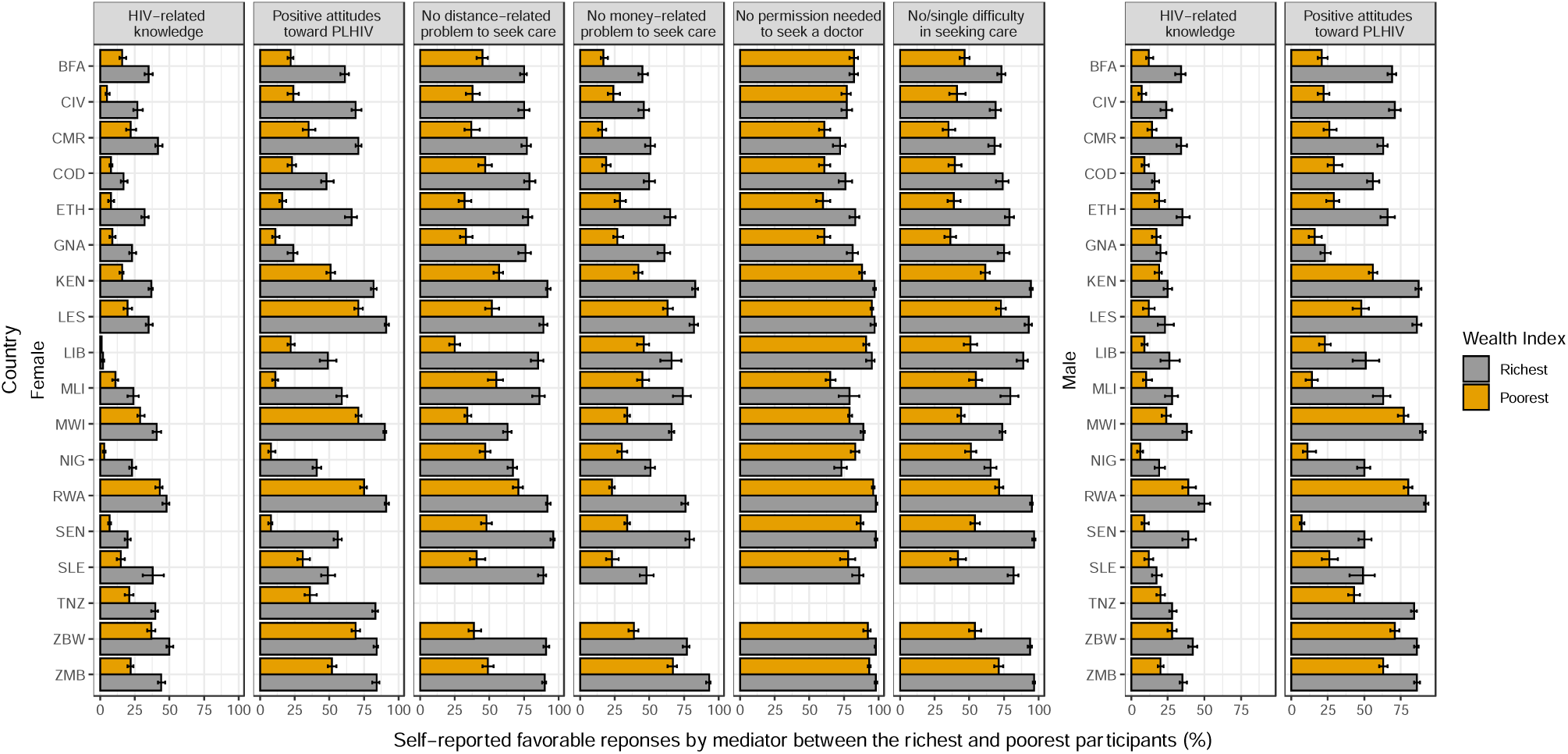
Path from exposure to mediator. Proportion of the richest and poorest participants who self-reported favourable levels of the mediator in 18 sub-Saharan African countries, stratified by gender. Refer to Table 1 for full country names.

Table 2 shows the total effect of wealth on recent HIV testing which was the effect estimate we used to measure wealth-related inequalities (Equation 1). We estimated the adjusted PRs of recent HIV testing between the richest and the poorest participants while accounting for confounders. Applying the cut-off of PR ≥ 1.5 led us to keep nine countries for women and 15 countries for men in our final mediation analyses.

**Table 2.** Total effect of wealth on recent HIV testing. Adjusted prevalence ratios of recent HIV testing between the richest and poorest participants (stratified by gender), while accounting for confounders. Bold fonts indicate that the model is statistically significant and eligible (PR ≥ 1.5), grey colours indicate that the model is significant but ineligible, and normal fonts indicate that the model is not significant. Refer to Table 1 for full country names.

| Adjusted PR (95% Confidence Interval) |  |  |
| --- | --- | --- |
| *P (recent HIV testing) = f(wealth, confounders)* |  |  |
| ISO Code † | Female | Male |
| **BFA** | **2.74 (2.09 – 3.58)** | **14.97 (9.12 – 24.57)** |
| **CIV** | **3.39 (2.53 – 4.52)** | **8.93 (5.27 – 15.15)** |
| **CMR** | **2.68 (2.33 – 3.07)** | **3.95 (3.23 – 4.83)** |
| **COD** | **12.14 (7.34 – 20.08)** | **15.30 (8.67 – 26.98)** |
| **ETH** | **3.97 (3.14 – 5.01)** | **3.89 (3.00 – 5.05)** |
| **GNA** | **10.63 (6.57 – 17.19)** | **11.27 (4.93 – 25.74)** |
| **KEN** | **1.30 (1.23 – 1.37)** | **1.59 (1.47 – 1.71)** |
| **LES** | **0.90 (0.82 – 0.99)** | **1.61 (1.30 – 2.00)** |
| **LIB** | 1.21 (0.99 – 1.47) | **2.92 (1.94 – 4.38)** |
| **MLI** | **11.17 (7.08 – 17.63)** | **6.16 (2.14 – 17.72)** |
| **MWI** | **1.10 (1.04 – 1.17)** | **1.13 (1.01 – 1.26)** |
| **NIG** | **4.82 (3.23 – 7.17)** | **46.04 (10.47 – 202.43)** |
| **RWA** | 1.08 (0.99 – 1.19) | 0.93 (0.80 – 1.07) |
| **SEN** | **1.62 (1.30 – 2.01)** | **3.08 (1.75 – 5.44)** |
| **SLE** | **1.35 (1.07 – 1.70)** | **2.58 (1.45 – 4.61)** |
| **TNZ** | **1.44 (1.27 – 1.65)** | **1.68 (1.42 – 1.99)** |
| **ZBW** | 1.13 (1.00 – 1.28) | **1.37 (1.16 – 1.63)** |
| **ZMB** | **1.13 (1.04 – 1.23)** | **1.51 (1.37 – 1.66)** |

Abbreviations: PR, Prevalence Ratio; P, probability; ISO, International Organization for Standardization.
† Refer to Table 1 for full country names.

Levels of wealth-related inequalities vary greatly by country and gender with pro-rich inequalities in HIV testing in most countries. Inequalities tended to be higher among men than women. Wealth-related inequalities were markedly observed in WCA countries. Among women, the highest wealth-related inequalities were in Congo DR where the prevalence of recent testing among the richest women was 12.14 (95% CI 7.34 – 20.08) times greater than among the poorest women. Meanwhile in men, the highest level of inequality was in Niger where the prevalence of recent testing among the richest men was 46.04 (10.47 – 202.43) times greater than among the poorest men.

### Mediated Effects

Pathways from exposure to each mediator based on Figure 1 were explored (Table S3). Among the eligible models (i.e., with sufficient levels of inequalities) in Table 2, we observed that wealth was associated with majority of the mediators except for HIV-related knowledge among men in Sierra Leone, positive attitudes toward PLHIV among men in Guinea and no permission from spouse/partner needed to seek a doctor in Burkina Faso, Côte d’Ivoire, Liberia and Niger among women (Table S3). The paths from each mediator to outcome were also explored (Figure S2 and Table S4). In all eligible countries except Lesotho, all mediators were positively associated with recent HIV testing (Table S4).

There was heterogeneity in the importance and role of each mediator in the pathway between wealth and recent testing across countries and gender groups (Figure 3). *Demand-related* mediators tended to have higher proportions mediated compared to *supply-related* mediators in women, with magnitudes varying across countries. For example, among women, the total effect of wealth on recent HIV testing uptake was mediated by positive attitudes toward PLHIV by 31.46% (95% CI 20.14%-53.37%) in Senegal, but only by 4.34% (−0.12%-8.78%) in Niger. In other words, we could also say that wealth-related inequality in testing among women in Senegal could be explained by positive attitudes toward PLHIV by 31.46% (95% CI 20.14%-53.37%).

**Figure 3.**
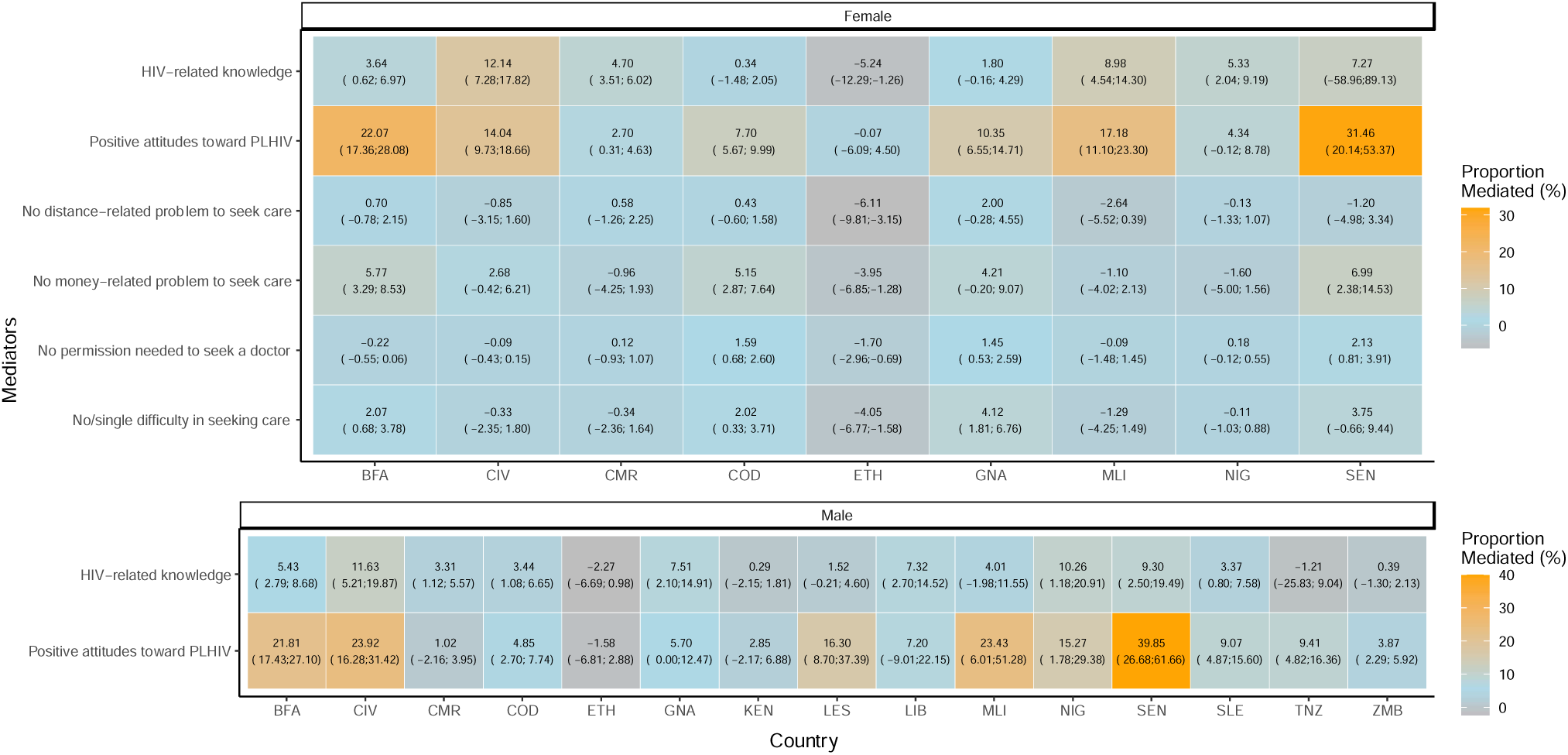
Heatmap of the proportion mediated by each mediator in the total effect of wealth on HIV testing, stratified by gender (eligible models). Refer to Table 1 for full country names.

Meanwhile, in Côte d’Ivoire, wealth-related inequalities in testing could be explained by HIV-related knowledge by 12.14% (7.28%-17.82%), while in Congo DR by only 0.34% (−1.48%-2.05%). *Supply-related* mediators tended to have lower proportions mediated in majority of the countries except in Burkina Faso, Congo DR, Guinea, and Senegal in which reporting no money-related problem mediated slightly more or almost similarly than HIV-related knowledge.

Among men, having positive attitudes toward PLHIV tended to mediate the total effect of wealth on testing more with a range between -1.58% (95CI -6.81%-2.88%) in Ethiopia and 39.85% (26.68%-61.66%) in Senegal than HIV-related knowledge that ranged between -2.27% (−6.69%-0.98%) in Ethiopia and 11.63% (5.21%-19.87%) in Côte d’Ivoire. A negative proportion mediated means that the CDE and NIE were in opposite direction. Figure S3 shows small reductions in wealth-related inequalities in a few countries after controlling for the mediators.

## Discussion

We analysed cross-sectional population-based surveys to assess the drivers of wealth-related inequalities in recent HIV testing uptake through mediation analysis in 18 SSA countries. Richest individuals were more likely to have been recently tested than the poorest with magnitudes varying across countries. We pre-identified several participant’s characteristics that could play a mediating role between wealth and recent uptake of HIV testing. The richest were more likely to have a favourable situation regarding these mediators (e.g., better knowledge about HIV, lesser stigma towards PLHIV and lesser problems to seek care) and these mediators were also positively associated with HIV testing. For instance, people having no problem to seek care were also more likely to have been recently tested for HIV. We found no single, strong mediator in the pathway between wealth and recent testing that was consistently strong across all countries and genders, but our results show that inequalities were mediated more by *demand*-(characterizes individual’s ability to perceive need for and inclination to seek health care) more than *supply-side* (ability to reach, pay for and engage in health care) characteristics. The importance of each mediator varied greatly by country and gender which may depend on several factors such as social, economic, epidemiology, donor and political structures of the country. This illustrates the importance of tailoring HIV testing programs to local context of the country and the needs of each gender.

Mediation analysis was conducted in countries where substantial levels of pro-rich wealth-related inequalities were observed, majority of which were WCA countries which is consistent to studies using different inequality estimates [6,25]. This is quite expected since most WCA countries’ health care delivery is through private sectors and often has inadequate decentralization of HIV services [26]. In ESA, on the other hand, health care delivery is mostly based on public and community health efforts incorporated with international donor funding [27]. Participants were also more likely to report having comprehensive HIV-related knowledge and positive attitudes toward PLHIV in ESA. This could be due to the longer history of HIV programs in this region in response to the higher burden of the epidemic. Inequalities were also found to be higher among men which could be explained by women having more access to HIV testing through routine offer in antenatal clinics as part of the prevention of mother-to-child transmission programs [28].

Countries with low uptake of recent testing tended to have high levels of inequalities with mediators having lower proportions mediated except for positive attitudes toward PLHIV. This may mean that attitudes toward PLHIV still plays a major role in explaining HIV testing inequalities regardless of the HIV epidemic in a country. HIV stigma has been well documented to be associated with higher SEP [29] and HIV testing uptake in the lifetime [30]. Stigma as an important mediator has implications for HIV testing. Due to the negative attitude towards PLHIV and the fear of being treated similarly, people may refuse to participate in any HIV prevention services or activities despite their knowledge [31]. It is also important to note that HIV-related knowledge and positive attitudes toward PLHIV do not influence testing uptake independently from each another based on an additional analysis using joint mediators approach (Table S5) [32]. In exploring the combined effects of the mediators, the mean of the individual specific PM should thus be considered instead of their sum for it will overestimate the combined proportions mediated of the mediators. This also applies to the *supply-related* mediators.

A study found that long travel times needed to reach healthcare in rural areas were found to be an important barrier in reaching 90% treatment coverage [33] and distance to care was found to affect uptake of facility delivery [34]. However, our findings showed that reporting no distance-related problem in seeking care mediated a lower proportion of the relationship between wealth and recent HIV testing uptake among women. We did not use physical distance itself but the perception that distance would be a problem in seeking care and in some countries like Senegal, HIV services reach the populations through both fixed and mobile strategies reinforced by mobile screening units [35]. Although magnitude is small, reporting no money-related problem in seeking care tended to have higher proportions mediated in WCA countries which have a widespread policy of user fees for health services [26]. In most countries, married women do not usually need spousal consent legally to access sexual and reproductive health facilities [36] making no permission needed from spouse/partner to seek a doctor mediate less.

The absence of a strong mediator that we could potentially control to reduce inequalities in recent testing across all countries and genders may be due to the fact these inequalities stem from country-level, rather than individual-level factors. Indeed a study found that upstream interventions such as structural interventions, provision of resources and fiscal interventions, among others, tended to reduce inequalities [37]. Meanwhile, downstream interventions such as media campaigns that focus only on individual factors like education were ineffective in reducing inequalities and were more likely to increase them [37].

This study has several limitations. First, the lack of a variable capturing risk perception of acquiring HIV in the DHS. A study found that risk perception is indeed an important mediator between peer education and HIV testing in key populations [38]. Second, the issue of temporality due to the cross-sectional nature of the data, especially for the *demand-related* mediators. Since counselling is part of HIV testing, we cannot exclude reverse causality between these *demand-related* mediators and HIV testing uptake. Another limitation is the self-report of HIV testing and mediators. A study, however, showed that the sensitivity of self-reported HIV testing ranged from 96% to 99% [39]. Despite this, reporting bias may still be present resulting to underreporting of sensitive information such as attitudes toward PLHIV. Another potential limitation is that inequalities have been measured only through wealth index which carries its own limits. Although asset-based wealth index is said to be stable and represents long-term SEP especially in LMIC, it can only assess relative wealth within a population [40]. For this reason, we did not pool the estimates across countries. Survey years were also different which may have contributed to the heterogeneity in inequality estimates and mediated effects.

Despite the limitations, this study has several strengths. We used large, standardized and nationally representative data. Moreover, to our knowledge, this is the first study to present a comprehensive dashboard of mediators in 18 sub-Saharan African countries. Importantly, compared to a classic mediation analysis, we used the potential outcomes framework allowing us to account for exposure-mediator interaction.

## Conclusions

Overall, the lack of an identified strong, single mediator illustrates that inequalities may not be addressed by solely acting upon a single factor but must be tackled upstream with social and structural interventions that address the root cause of the problem. Although, we were not able to identify a single strong mediator, we were able to underline the use of mediation analysis based on the potential outcomes framework in assessing socioeconomic inequalities in HIV testing.

More research is needed to explore other potential mediators and contextual factors. Beyond measuring inequalities in HIV testing, we need to understand the determinants of inequalities in HIV testing to direct specific interventions that could reduce inequalities and meet the needs of everyone.

## Data Availability

Raw data from the DHS surveys used in this study are publicly available for academic research (www.dhsprogram.com). Formatted and processed data supporting the findings of this study are available from the corresponding author on request.

## Authors’ contributions

PAAT, LT and KJ conceived and discussed the study with input from TB. PAAT and MH collated and processed the DHS. PAAT conducted the analysis with inputs from TB, LT and KJ. PAAT produced output figures and tables with inputs from LT and KJ. All authors contributed to the interpretation of the results. PAAT wrote the initial draft with inputs from LT and KJ. All authors contributed to subsequent revisions.

## Acknowledgements

This study was funded by INSERM-ANRS (France Recherche Nord and Sud Sida-HIV Hépatites), grant number ANRS 12377-B104.

## Disclaimer

Funding agency had no role in the study design, data collection and analysis.

## Additional files

Additional file 1: **Supplementary material**

Description:

**Figure S1**. Categorization of the mediators.

**Table S1**. Construction and coding of the mediators.

**Text S1**. Formulas based on Valeri and Vanderweele to estimate the Control Direct Effect. Natural Indirect Effect, Total Effect and Proportion mediated.

**Table S2**. Survey and population characteristics, by country and gender.

**Table S3**. Path from exposure to mediator. Adjusted prevalence ratios of favorable levels of the mediator between the richest and poorest participants while accounting for confounders.

**Figure S2**. Path from mediator to outcome. Proportion of HIV testing uptake among the favorable and unfavorable levels of the mediator in 18 sub-Saharan African countries, stratified by gender.

**Table S4**. Path from mediator to outcome. Adjusted prevalence ratios of recent HIV testing between favorable and unfavorable levels of the mediators, while accounting for confounders.

**Figure S3**. Forest plot of the Total Effect and Controlled Direct Effect by mediator and gender.

**Table S5**. Proportion mediated by individual and joint mediators, stratified by gender, in 18 sub-Saharan African countries.

