## Supplementary Materials for "Understanding the pathways leading to socioeconomic inequalities in HIV testing uptake in 18 sub-Saharan African countries: a mediation analysis"

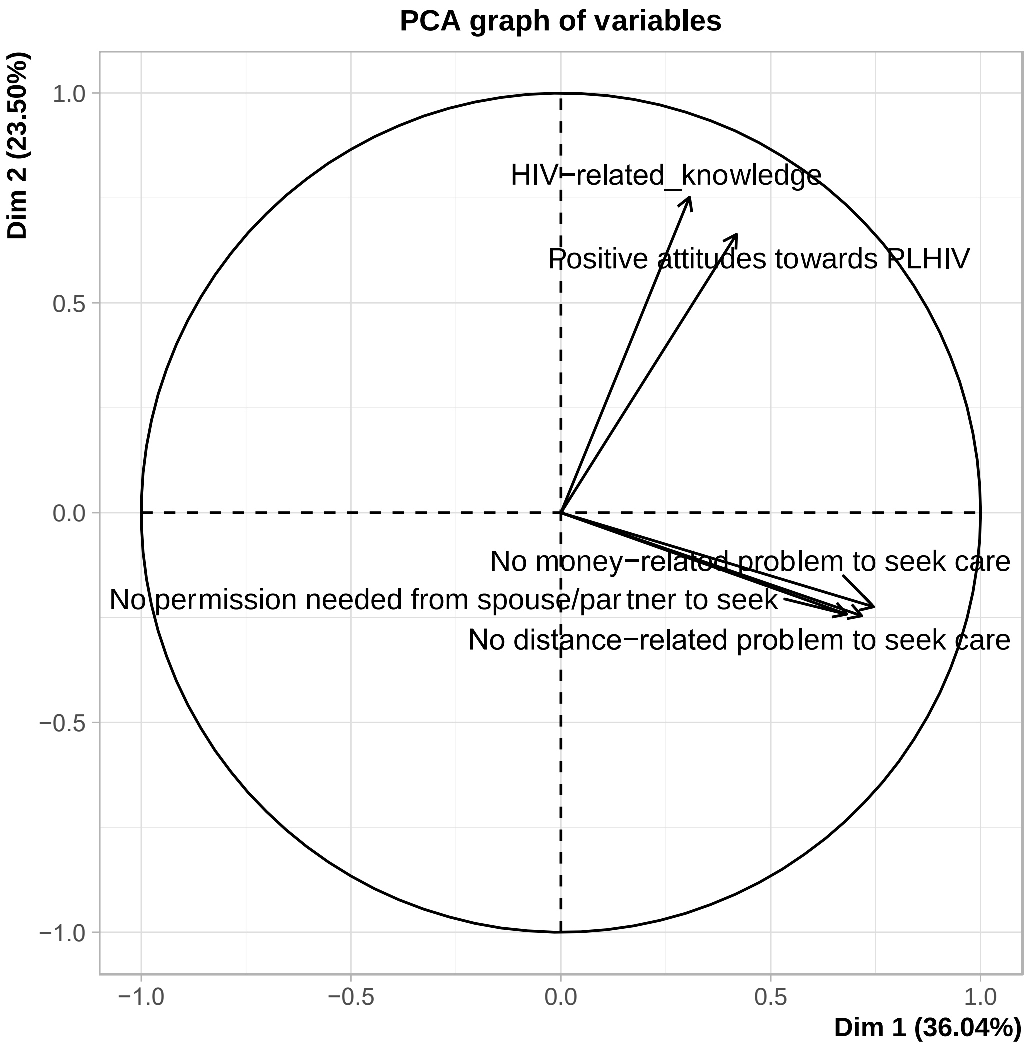

**Figure S1. Categorization of the mediators.** Principal Component Analysis of the six mediators among female participants in 18 sub-Saharan African countries.

Among women, the PCA show the *demand-related* mediators were highly correlated. We found the same result across the *supply-related* mediators. Hence, we categorized these mediators under these two groups.

Among men, *demand-related* mediators were also highly correlated (Pearson chi-square p-value: < 2.2e-16).

### Table S1. Construction and coding of the mediators.

| Variables | Description | Coding | DHS Questions |
|---|---|---|---|
| **Mediators** | | | |
| **Demand-related mediators** | | | |
| **HIV-related knowledge** | Based on a set of 7 questions related to HIV transmission and prevention defined as a binary variable reflecting comprehensive knowledge about HIV | 1 = answering correctly to 7 questions<br><br>0 = answering at least one incorrectly | 1. Can people reduce their chance of getting HIV by having just one uninfected sex partner who has no other sex partners?<br>2. Can people get HIV from mosquito bites?<br>3. Can people reduce their chance of getting HIV by using a condom every time they have sex?<br>4. Can people get HIV by sharing food with a person who has HIV?<br>5. Can HIV be transmitted from a mother to her baby:<br>a. During delivery?<br>b. During pregnancy?<br>c. By breastfeeding? |
| **Positive attitudes towards PLHIV** | Based on a set of 2 questions about attitudes towards PLHIV defined as a binary variable showing positive attitudes towards PLHIV. The set of two questions differ by country depending on the availability of the variables. | 1 = answering favorably to 2 questions<br><br>0 = answering at least one unfavorably | 1. Would want to keep secret that a family member got infected with HIV<br>2. Willing to care for a family member with the AIDS virus<br>3. Female teacher who has the AIDS virus but is not sick should be allowed to continue teaching<br>4. Would buy vegetables from HIV-positive-vendor<br>5. Children with HIV+ should not be allowed to go to the same school |
| **Supply-related mediators (among females only)** | | | Many different factors can prevent women from getting medical advice or treatment for themselves. When you are sick and want to get medical advice or treatment, is each of the following a big problem or not a big problem: |
| **Reporting no distance-related problem to seek care** | Self-report of distance to facility as not a problem/ not a big problem. | 1 = yes<br><br>0 = no | 1. The distance to the health facility? |
| **Reporting no money-related problem to seek care** | Self-report of getting money for medical advice or treatment as not a problem/ not a big problem. | 1 = yes<br><br>0 = no | 2. Getting money needed for advice or treatment? |
| **No permission needed from spouse/partner to seek a doctor** | Self-report of getting permission to see a doctor as not a problem/ not a big problem. | 1 = yes<br><br>0 = no | 3. Getting permission to go to the doctor? |
| **No/ single difficulty in seeking care** | Only one or no difficulty in access. Combined variable of the 3 proxy variables related to offer. | 1 = yes<br><br>0 = no | |

**Text S1. Formulas based on Valeri and Vanderweele to estimate the Control Direct Effect (CDE), Natural Indirect Effect (NIE), Total Effect (TE) and Proportion mediated (PM).**

$$\mathrm{E}\{(Y = 1|a, m, c)\} = \theta o + \theta 1a + \theta 2m + \theta 3am + \theta' 4c \quad \text{(Outcome model)}$$

$$\mathrm{E}\{(M = 1|a, c)\} = \beta o + \beta 1a + \beta' 2c \quad \text{(Mediator model)}$$

where, $Y$ = outcome, $a$ = exposure, $m$ = mediator, $c$ = confounder.

$$PR^{CDE} = \exp\{(\theta 1 + \theta 3m)(a - a*)\}, \text{ m=1}$$

$$PR^{NIE} = \frac{\{1 + \exp(\beta 0 + \beta 1a * + \beta' 2c)\}\{1 + \exp(\theta 2 + \theta 3a + \beta 0 + \beta 1a + \beta' 2c)\}}{\{1 + \exp(\beta 0 + \beta 1a + \beta' 2c)\}\{1 + \exp(\theta 2 + \theta 3a + \beta 0 + \beta 1a * + \beta' 2c)\}}$$

where $a$ = 1, $a$* = 0, $m$ = 1.

TE = NIE x CDE (when Y is binary)

$$PM = CDE\ x\ (NIE - 1)/(CDE\ x\ NIE - 1)$$

Valeri L, VanderWeele TJ. "Mediation analysis allowing for exposure–mediator interactions and causal interpretation: Theoretical assumptions and implementation with SAS and SPSS macros": Correction to Valeri and VanderWeele (2013). Psychol Methods. 2013 Dec;18(4):474–474.

### Table S2. Survey and population characteristics, by country and gender.

| Western-Central Africa | BFA (Burkina Faso) 2010 | | CIV (Côte d'Ivoire) 2011-12 | | CMR (Cameroon) 2018 | | COD (Congo DR) 2013-14 | | GNA (Guinea) 2018 | | LIB (Liberia) 2013 | | MLI (Mali) 2018 | | NIG (Niger) 2012 | | SEN (Senegal) 2017 | | SLE (Sierra Leone) 2013 | |
|---|---|---|---|---|---|---|---|---|---|---|---|---|---|---|---|---|---|---|---|---|
| | Female | Male | Female | Male | Female | Male | Female | Male | Female | Male | Female | Male | Female | Male | Female | Male | Female | Male | Female | Male |
| N | 17,087 | 7,307 | 10,060 | 5,135 | 14,677 | 6,978 | 18,827 | 8,656 | 10,874 | 4,117 | 9,239 | 4,118 | 10,519 | 4,618 | 11,160 | 3,928 | 16,787 | 6,977 | 16,658 | 7,262 |
| Response Rate (%) * | 98 | 97 | 93 | 91 | 98 | 98 | 99 | 97 | 99 | 97 | 98 | 95 | 98 | 96 | 95 | 88 | 96 | 91 | 97 | 96 |
| Wealth index (%) Poorest | 17.50 | 17.30 | 17.60 | 19.10 | 16.60 | 15.40 | 18.60 | 16.90 | 18.90 | 17.30 | 17.10 | 18.20 | 17.50 | 18.60 | 18.10 | 14.80 | 16.50 | 16.70 | 18.50 | 18.50 |
| Poorer | 18.70 | 19.10 | 17.30 | 16.50 | 18.70 | 18.40 | 19.10 | 18.90 | 19.70 | 18.00 | 17.60 | 18.30 | 18.80 | 19.90 | 18.80 | 17.40 | 17.80 | 17.50 | 18.30 | 18.30 |
| Middle | 19.00 | 18.30 | 18.20 | 17.90 | 20.10 | 20.80 | 18.60 | 20.70 | 18.90 | 18.00 | 19.30 | 17.70 | 19.10 | 18.60 | 19.70 | 19.30 | 19.70 | 19.80 | 18.80 | 18.50 |
| Richer | 19.90 | 18.90 | 20.80 | 22.10 | 21.30 | 21.00 | 19.40 | 20.50 | 19.80 | 20.30 | 22.20 | 21.00 | 21.10 | 20.10 | 20.60 | 20.40 | 21.30 | 22.80 | 20.30 | 18.10 |
| Richest | 24.90 | 26.40 | 26.20 | 24.40 | 23.20 | 24.30 | 24.30 | 23.00 | 22.70 | 26.40 | 23.90 | 24.90 | 23.40 | 22.80 | 22.80 | 28.10 | 24.70 | 23.20 | 24.00 | 26.50 |
| Age in years (%) 15-24 | 38.80 | 33.80 | 39.50 | 33.90 | 39.00 | 38.20 | 41.20 | 36.40 | 40.10 | 35.90 | 40.30 | 38.50 | 38.00 | 31.20 | 34.20 | 28.00 | 40.60 | 39.90 | 39.40 | 34.20 |
| 25-34 | 32.50 | 25.90 | 34.10 | 29.40 | 30.00 | 25.40 | 32.70 | 26.40 | 30.80 | 23.00 | 30.40 | 30.30 | 34.40 | 24.60 | 37.00 | 24.80 | 32.20 | 25.50 | 30.80 | 25.10 |
| 35 and above | 28.80 | 40.30 | 26.40 | 36.80 | 31.00 | 36.50 | 26.10 | 37.20 | 29.10 | 41.10 | 29.30 | 31.20 | 27.60 | 44.20 | 28.80 | 47.20 | 27.30 | 34.60 | 29.80 | 40.80 |
| Type of residence (%) Urban | 27.10 | 28.90 | 51.40 | 50.30 | 54.60 | 55.20 | 38.40 | 37.00 | 37.60 | 41.90 | 61.00 | 58.60 | 26.30 | 25.60 | 18.80 | 24.60 | 49.70 | 53.10 | 35.60 | 37.20 |
| Family situation (%) In union | 79.40 | 63.70 | 62.70 | 52.70 | 56.60 | 47.10 | 64.20 | 58.20 | 71.10 | 55.10 | 58.30 | 53.90 | 81.40 | 66.10 | 88.50 | 69.80 | 64.90 | 43.10 | 65.50 | 57.10 |
| Single | 17.50 | 34.20 | 30.20 | 42.40 | 32.30 | 48.20 | 26.00 | 37.50 | 25.20 | 43.60 | 31.00 | 42.50 | 16.00 | 33.10 | 7.90 | 28.60 | 30.30 | 55.50 | 28.40 | 39.30 |
| Widowed/ separated | 3.10 | 2.20 | 7.10 | 4.90 | 11.10 | 4.80 | 9.70 | 4.20 | 3.70 | 1.30 | 10.70 | 3.70 | 2.60 | 0.80 | 3.60 | 1.50 | 4.80 | 1.40 | 6.20 | 3.50 |

† Based on each country's DHS Final Report

‡ Women: 31,079 (15-49 years of age: all version)
Women: 14,741 (15-49 years of age: full version); 96%
Women: 16,338 (15-49 years of age: short version); 97%

### Table S2 (continued). Survey and population characteristics, by country and gender.

| Eastern-Southern Africa | ETH (Ethiopia) | | KEN (Kenya) | | LES (Lesotho) | | MWI (Malawi) | | RWA (Rwanda) | | TNZ (Tanzania) | | ZBW (Zimbabwe) | | ZMB (Zambia) | |
|---|---|---|---|---|---|---|---|---|---|---|---|---|---|---|---|---|
| | 2016 | | 2014 | | 2014 | | 2015-16 | | 2014-15 | | 2011-12 | | 2015 | | 2018 | |
| | Female | Male | Female‡ | Male | Female | Male | Female | Male | Female | Male | Female | Male | Female | Male | Female | Male |
| N | 15 683 | 12 688 | 31 079 | 12 819 | 6 621 | 2 931 | 24 562 | 7 478 | 13 497 | 6 217 | 10 967 | 8 352 | 9 955 | 8 396 | 13 683 | 12 132 |
| Response Rate (%) † | 95 | 86 | 97 | 90 | 97 | 94 | 98 | 95 | 99.5 | 99.5 | 96 | 89 | 96 | 92 | 96 | 92 |
| Wealth index (%) Poorest | 16.80 | 15.80 | 15.60 | 14.10 | 14.50 | 14.30 | 19.30 | 16.00 | 19.00 | 14.60 | 17.00 | 16.30 | 17.10 | 15.00 | 17.80 | 16.50 |
| Poorer | 17.90 | 18.30 | 17.60 | 17.70 | 15.60 | 18.20 | 19.10 | 18.40 | 19.50 | 17.80 | 18.00 | 18.30 | 17.00 | 18.00 | 17.40 | 17.90 |
| Middle | 19.00 | 19.30 | 19.40 | 19.80 | 18.80 | 20.40 | 18.90 | 19.80 | 19.20 | 20.20 | 18.00 | 19.00 | 17.60 | 19.30 | 18.10 | 20.00 |
| Richer | 19.80 | 21.50 | 21.10 | 24.60 | 24.20 | 22.50 | 19.10 | 20.70 | 19.50 | 22.70 | 20.60 | 20.90 | 23.20 | 22.90 | 22.00 | 22.40 |
| Richest | 26.50 | 25.10 | 26.40 | 23.90 | 26.90 | 24.60 | 23.70 | 25.20 | 22.80 | 24.70 | 26.40 | 25.40 | 25.10 | 24.70 | 24.60 | 23.30 |
| Age in years (%) 15-24 | 39.20 | 35.10 | 37.20 | 36.40 | 41.80 | 42.70 | 42.40 | 43.10 | 38.70 | 36.60 | 39.20 | 42.30 | 39.10 | 41.20 | 41.90 | 39.70 |
| 25-34 | 33.80 | 28.50 | 34.10 | 30.30 | 31.00 | 25.40 | 31.00 | 26.00 | 33.00 | 30.20 | 31.00 | 26.10 | 32.90 | 27.00 | 30.00 | 25.60 |
| 35 and above | 27.00 | 36.40 | 28.70 | 33.30 | 27.30 | 31.90 | 26.50 | 30.80 | 28.30 | 33.20 | 29.80 | 31.50 | 28.00 | 31.80 | 28.10 | 34.70 |
| Type of residence (%) Urban | 22.20 | 19.70 | 40.80 | 43.40 | 36.50 | 33.80 | 18.30 | 18.50 | 19.50 | 20.00 | 27.00 | 25.60 | 38.50 | 36.00 | 46.60 | 44.10 |
| Family situation (%) In union | 65.20 | 58.90 | 59.70 | 52.70 | 54.60 | 40.00 | 65.70 | 58.10 | 51.70 | 54.20 | 63.00 | 53.00 | 61.80 | 51.50 | 55.90 | 53.00 |
| Single | 25.70 | 38.60 | 28.90 | 41.80 | 33.10 | 51.50 | 21.00 | 38.30 | 37.80 | 43.50 | 25.50 | 42.30 | 25.20 | 43.20 | 31.20 | 42.60 |
| Widowed/ separated | 9.10 | 2.50 | 11.40 | 5.40 | 12.40 | 8.50 | 13.30 | 3.50 | 10.50 | 2.30 | 11.50 | 4.70 | 13.00 | 5.30 | 12.90 | 4.40 |

† Based on each country's DHS Final Report

‡ Women: 31,079 (15-49 years of age: all version)
Women: 14,741 (15-49 years of age: full version); 96%
Women: 16,338 (15-49 years of age: short version); 97%

### Table S3. Path from exposure to mediator. Adjusted prevalence ratios of favorable levels of the mediator between the richest and poorest participants while accounting for confounders.

| Country ‡ | Adjusted PR (95% Confidence Intervals) † $P\ (mediator) = f(wealth, confounders)$ Mediator | | | | | | | |
|---|---|---|---|---|---|---|---|---|
| | HIV-related knowledge | | Positive attitudes toward PLHIV | | No distance-related problem to seek care | No money-related problem to seek care | No permission needed to seek a doctor | No/single difficulty in seeking care |
| | Female | Male | Female | Male | Female | Female | Female | Female |
| **BFA** | **2.04 [1.68;2.46]** | **2.46 [1.87;3.24]** | **2.43 [2.05;2.89]** | **2.98 [2.40;3.71]** | **1.56 [1.38;1.77]** | **3.68 [2.95;4.58]** | 0.93 [0.85;1.01] | **1.52 [1.35;1.72]** |
| **CIV** | **5.20 [3.75;7.21]** | **5.99 [3.92;9.16]** | **3.18 [2.68;3.76]** | **4.87 [4.05;5.85]** | **2.01 [1.72;2.34]** | **3.99 [3.20;4.98]** | 1.06 [0.95;1.19] | **2.06 [1.75;2.42]** |
| **CMR** | **1.95 [1.66;2.30]** | **2.36 [1.81;3.09]** | **1.96 [1.76;2.19]** | **2.18 [1.85;2.57]** | **1.89 [1.69;2.11]** | **4.71 [3.91;5.68]** | **1.38 [1.24;1.55]** | **2.06 [1.80;2.37]** |
| **COD** | **2.05 [1.55;2.70]** | **2.29 [1.56;3.37]** | **1.93 [1.57;2.37]** | **1.73 [1.42;2.12]** | **1.46 [1.27;1.69]** | **3.20 [2.53;4.03]** | **1.35 [1.22;1.49]** | **1.82 [1.56;2.13]** |
| **ETH** | **6.33 [4.83;8.30]** | **3.36 [2.63;4.30]** | **4.00 [3.27;4.90]** | **2.99 [2.45;3.64]** | **2.46 [2.06;2.94]** | **3.00 [2.55;3.54]** | **1.32 [1.20;1.46]** | **2.07 [1.79;2.39]** |
| **GNA** | **1.82 [1.25;2.65]** | **1.75 [1.07;2.84]** | **2.69 [1.71;4.23]** | 1.49 [0.82;2.72] | **2.16 [1.76;2.64]** | **3.14 [2.50;3.96]** | **1.25 [1.10;1.42]** | **2.10 [1.75;2.53]** |
| **KEN** | **2.69 [2.37;3.05]** | **1.61 [1.39;1.87]** | **2.04 [1.91;2.18]** | **1.77 [1.68;1.88]** | **1.73 [1.63;1.84]** | **2.33 [2.16;2.50]** | **1.13 [1.10;1.16]** | **1.68 [1.59;1.76]** |
| **LES** | **2.14[1.76;2.60]** | **2.12 [1.42;3.17]** | **1.34 [1.27;1.42]** | **1.96 [1.74;2.20]** | **1.75[1.58;1.93]** | **1.45 [1.34;1.56]** | **1.03 [1.01;1.05]** | **1.32 [1.25;1.40]** |
| **LIB** | **3.93[1.42;10.89]** | **2.58 [1.68;3.95]** | **2.76 [2.20;3.45]** | **2.69 [2.06;3.51]** | **2.94 [2.31;3.73]** | **1.39 [1.20;1.61]** | 1.02 [0.98;1.07] | **1.61 [1.41;1.84]** |

Abbreviations: PR, prevalence ratio; P, probability.

† Legend: **Statistically significant,** Not statistically significant

‡ Refer to Table S2 for full country names

**Table S3 (continued). Path from exposure to mediator. Adjusted prevalence rations of favorable levels of the mediator between the richest and poorest participants while accounting for confounders.**

Adjusted PR (95% Confidence Intervals) †

$$P\ (mediator) = f(wealth, confounders)$$

| Country ‡ | Mediator | | | | | | | |
|---|---|---|---|---|---|---|---|---|
| | HIV-related knowledge | | Positive attitudes toward PLHIV | | No distance-related problem to seek care | No money-related problem to seek care | No permission needed to seek a doctor | No/single difficulty in seeking care |
| | Female | Male | Female | Male | Female | Female | Female | Female |
| MLI | 5.06 [3.34;7.66] | 3.67 [2.31;5.84] | 10.15 [7.34;14.04] | 8.63 [6.30;11.83] | 2.16 [1.88;2.48] | 2.68 [2.28;3.14] | 1.45 [1.30;1.61] | 2.17 [1.91;2.47] |
| MWI | 1.56 [1.43;1.70] | 1.60 [1.36;1.89] | 1.31 [1.27;1.35] | 1.21 [1.16;1.26] | 1.92 [1.74;2.12] | 2.37 [2.20;2.55] | 1.15 [1.12;1.19] | 1.87 [1.76;2.00] |
| NIG | 6.42 [4.11;10.03] | 3.69 [2.01;6.77] | 5.50 [3.61;8.36] | 6.49 [4.09;10.29] | 1.45 [1.25;1.69] | 2.16 [1.77;2.64] | 0.92 [0.83;1.03] | 1.35 [1.16;1.57] |
| RWA | 1.08 [0.98;1.18] | 1.10 [0.92;1.32] | 1.23 [1.19;1.28] | 1.19 [1.14;1.26] | 1.26 [1.19;1.33] | 4.70 [4.21;5.25] | 1.03 [1.02;1.05] | 1.38 [1.31;1.44] |
| SEN | 3.58 [2.89;4.44] | 3.88 [2.79;5.39] | 6.90 [5.90;8.06] | 6.42 [5.14;8.01] | 2.01 [1.84;2.20] | 3.25 [2.97;3.56] | 1.11 [1.08;1.14] | 1.88 [1.75;2.02] |
| SLE | 2.11 [1.52;2.94] | 1.72 [0.95;3.10] | 1.48 [1.19;1.83] | 1.76 [1.23;2.52] | 1.89 [1.61;2.23] | 2.74 [1.94;3.85] | 1.16 [1.05;1.27] | 1.94 [1.61;2.32] |
| TNZ | 1.68 [1.45;1.94] | 1.34 [1.13;1.60] | 2.25 [2.05;2.47] | 2.11 [1.93;2.31] | NA | NA | NA | NA |
| ZBW | 1.61 [1.40;1.85] | 1.70 [1.42;2.05] | 1.32 [1.23;1.42] | 1.34 [1.24;1.44] | 1.99 [1.76;2.25] | 3.02 [2.67;3.41] | 1.09 [1.06;1.13] | 1.79 [1.64;1.95] |
| ZMB | 2.33 [2.01;2.69] | 2.22 [1.86;2.66] | 1.91 [1.78;2.06] | 1.59 [1.49;1.70] | 1.69 [1.51;1.90] | 1.43 [1.33;1.53] | 1.06 [1.03;1.08] | 1.35 [1.26;1.43] |

Abbreviations: PR, prevalence ratio; P, probability.

† Legend: **Statistically significant,** Not statistically significant

‡ Refer to Table S2 for full country names

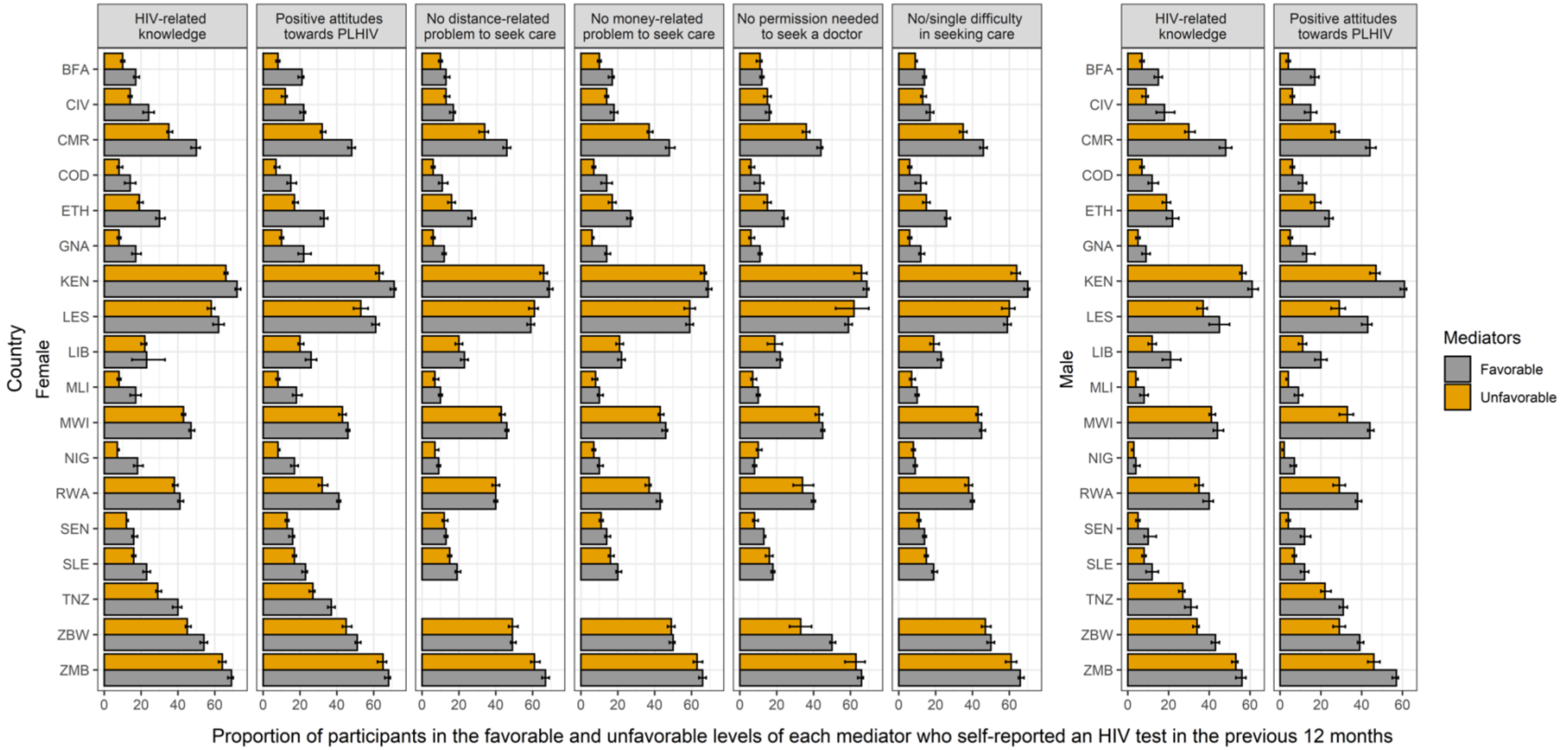

**Figure S2. Path from mediator to outcome. Bivariate analysis of HIV testing uptake and mediators. Proportion of HIV testing uptake among the favorable and unfavorable levels of the mediator in 18 sub-Saharan African countries, stratified by gender**. Refer to Table S2 for full country names.

### Table S4. Path from mediator to outcome. Adjusted prevalence ratios of recent HIV testing between favorable and unfavorable levels of the mediators, while accounting for confounders.

| | | Adjusted PR (95% Confidence Intervals) † $P\ (recent\ HIV\ testing) = f(mediator, confounders)$ | | | | | |
|---|---|---|---|---|---|---|---|
| Country † | Mediators | HIV-related knowledge | Positive attitudes toward PLHIV | No distance-related problem to seek care | No money-related problem to seek care | No permission needed to seek a doctor | No/ single difficulty in seeking care |
| BFA | Female | **1.42 [1.29;1.57]** | **2.09 [1.88;2.32]** | 1.10 [0.98;1.23] | **1.36 [1.23;1.51]** | 1.07 [0.94;1.22] | **1.20 [1.08;1.35]** |
| | Male | **1.56 [1.35;1.82]** | **2.90 [2.41;3.49]** | NA | NA | NA | NA |
| CIV | Female | **1.51 [1.35;1.69]** | **1.58 [1.40;1.77]** | 1.03 [0.91;1.16] | **1.18 [1.06;1.32]** | 0.97 [0.84;1.11] | 1.05 [0.94;1.18] |
| | Male | **1.82 [1.50;2.21]** | **2.27 [1.86;2.76]** | NA | NA | NA | NA |
| CMR | Female | **1.29 [1.24;1.35]** | **1.33 [1.26;1.41]** | **1.20 [1.13;1.27]** | **1.19 [1.13;1.25]** | **1.18 [1.11;1.25]** | **1.17 [1.11;1.24]** |
| | Male | **1.46 [1.36;1.57]** | **1.55 [1.41;1.70]** | NA | NA | NA | NA |
| COD | Female | **1.38 [1.21;1.57]** | **1.79 [1.58;2.02]** | **1.17 [1.01;1.35]** | **1.54 [1.35;1.75]** | **1.34 [1.15;1.56]** | **1.32 [1.14;1.54]** |
| | Male | **1.52 [1.25;1.86]** | **1.67 [1.37;2.05]** | NA | NA | NA | NA |
| ETH | Female | **1.21 [1.13;1.29]** | **1.54 [1.42;1.65]** | **1.16 [1.07;1.26]** | **1.20 [1.12;1.28]** | **1.14 [1.05;1.23]** | **1.19 [1.10;1.28]** |
| | Male | **1.13 [1.05;1.22]** | **1.37 [1.25;1.50]** | NA | NA | NA | NA |
| GNA | Female | **1.56 [1.35;1.80]** | **2.07 [1.73;2.46]** | **1.25 [1.05;1.49]** | **1.76 [1.52;2.04]** | **1.35 [1.10;1.65]** | **1.41 [1.18;1.69]** |
| | Male | 1.36 [1.00;1.84] | **2.33 [1.75;3.09]** | NA | NA | NA | NA |
| KEN | Female | **1.10 [1.08;1.13]** | **1.19 [1.15;1.22]** | **1.09 [1.06;1.13]** | **1.04 [1.02;1.07]** | **1.06 [1.01;1.12]** | **1.13 [1.09;1.17]** |
| | Male | **1.12 [1.08;1.16]** | **1.35 [1.29;1.42]** | NA | NA | NA | NA |
| LES | Female | 1.04 [1.00;1.09] | **1.11 [1.05;1.18]** | 1.02 [0.98;1.07] | 1.01 [0.97;1.06] | 0.95 [0.86;1.05] | 1.01 [0.96;1.06] |
| | Male | **1.13 [1.02;1.26]** | **1.45 [1.29;1.62]** | NA | NA | NA | NA |
| LIB | Female | 1.18 [0.90;1.55] | **1.29 [1.18;1.42]** | **1.12 [1.01;1.23]** | 1.10 [1.00;1.21] | **1.27 [1.09;1.48]** | **1.14 [1.03;1.26]** |
| | Male | **1.51 [1.25;1.82]** | **1.61 [1.36;1.91]** | NA | NA | NA | NA |

Abbreviations: PR, prevalence ratio; P, probability.

† Legend: **Statistically significant,** Not statistically significant

‡ Refer to Table S2 for full country names

**Table S4 (continued). Path from mediator to outcome. Adjusted prevalence ratios of recent HIV testing between favorable and unfavorable levels of the mediators, while accounting for confounders.**

| | | Adjusted PR (95% Confidence Intervals) † $P\ (recent\ HIV\ testing) = f(mediator, confounders)$ | | | | | |
|---|---|---|---|---|---|---|---|
| Country ‡ | Mediators | HIV-related knowledge | Positive attitudes toward PLHIV | No distance-related problem to seek care | No money-related problem to seek care | No permission needed to seek a doctor | No/ single difficulty to in seeking care |
| **MLI** | **Female** | **1.63 [1.34;1.98]** | **1.91 [1.60;2.28]** | 1.14 [0.93;1.41] | **1.21 [1.01;1.45]** | 1.14 [0.89;1.47] | 1.19 [0.97;1.46] |
| | **Male** | **1.33 [1.01;1.75]** | **1.58 [1.12;2.22]** | NA | NA | NA | NA |
| **MWI** | **Female** | **1.05 [1.01;1.08]** | **1.05 [1.01;1.09]** | 1.02 [0.99;1.05] | **1.04 [1.01;1.07]** | 1.04 [0.99;1.08] | 1.02 [0.99;1.05] |
| | **Male** | 1.04 [0.98;1.10] | **1.17 [1.08;1.27]** | NA | NA | NA | NA |
| **NIG** | **Female** | **1.36 [1.17;1.58]** | **1.24 [1.10;1.40]** | 1.02 [0.89;1.17] | **1.18 [1.04;1.33]** | 0.91 [0.80;1.05] | 1.02 [0.89;1.15] |
| | **Male** | **1.61 [1.10;2.37]** | **1.86 [1.29;2.68]** | NA | NA | NA | NA |
| **RWA** | **Female** | **1.06 [1.02;1.11]** | **1.22 [1.14;1.30]** | 0.99 [0.93;1.04] | **1.13 [1.08;1.17]** | 1.15 [0.99;1.35] | 1.04 [0.98;1.11] |
| | **Male** | **1.10 [1.03;1.18]** | **1.28 [1.15;1.43]** | NA | NA | NA | NA |
| **SEN** | **Female** | **1.26 [1.14;1.40]** | **1.42 [1.30;1.55]** | 1.05 [0.94;1.17] | **1.20 [1.10;1.31]** | **1.40 [1.16;1.69]** | **1.17 [1.04;1.31]** |
| | **Male** | **1.48 [1.15;1.89]** | **2.23 [1.81;2.75]** | NA | NA | NA | NA |
| **SLE** | **Female** | **1.30 [1.18;1.42]** | **1.38 [1.27;1.51]** | **1.12 [1.02;1.24]** | **1.20 [1.10;1.31]** | 1.09 [0.97;1.21] | **1.12 [1.02;1.22]** |
| | **Male** | **1.39 [1.12;1.72]** | **1.67 [1.35;2.07]** | NA | NA | NA | NA |
| **TNZ** | **Female** | **1.34 [1.26;1.42]** | **1.31 [1.23;1.39]** | NA | NA | NA | NA |
| | **Male** | **1.14 [1.05;1.24]** | **1.28 [1.17;1.39]** | NA | NA | NA | NA |
| **ZBW** | **Female** | **1.09 [1.05;1.14]** | **1.10 [1.05;1.16]** | 1.04 [0.99;1.09] | **1.07 [1.02;1.11]** | **1.43 [1.25;1.63]** | **1.09 [1.04;1.15]** |
| | **Male** | **1.15 [1.09;1.22]** | **1.18 [1.09;1.27]** | NA | NA | NA | NA |
| **ZMB** | **Female** | **1.04 [1.01;1.07]** | **1.06 [1.03;1.09]** | **1.07 [1.03;1.11]** | 1.02 [0.98;1.06] | 1.00 [0.93;1.08] | 1.05 [1.00;1.09] |
| | **Male** | 1.03 [0.99;1.07] | **1.15 [1.10;1.20]** | NA | NA | NA | NA |

Abbreviations: PR, prevalence ratio; P, probability.

† Legend: **Statistically significant,** Not statistically significant

‡ Refer to Table S2 for full country names

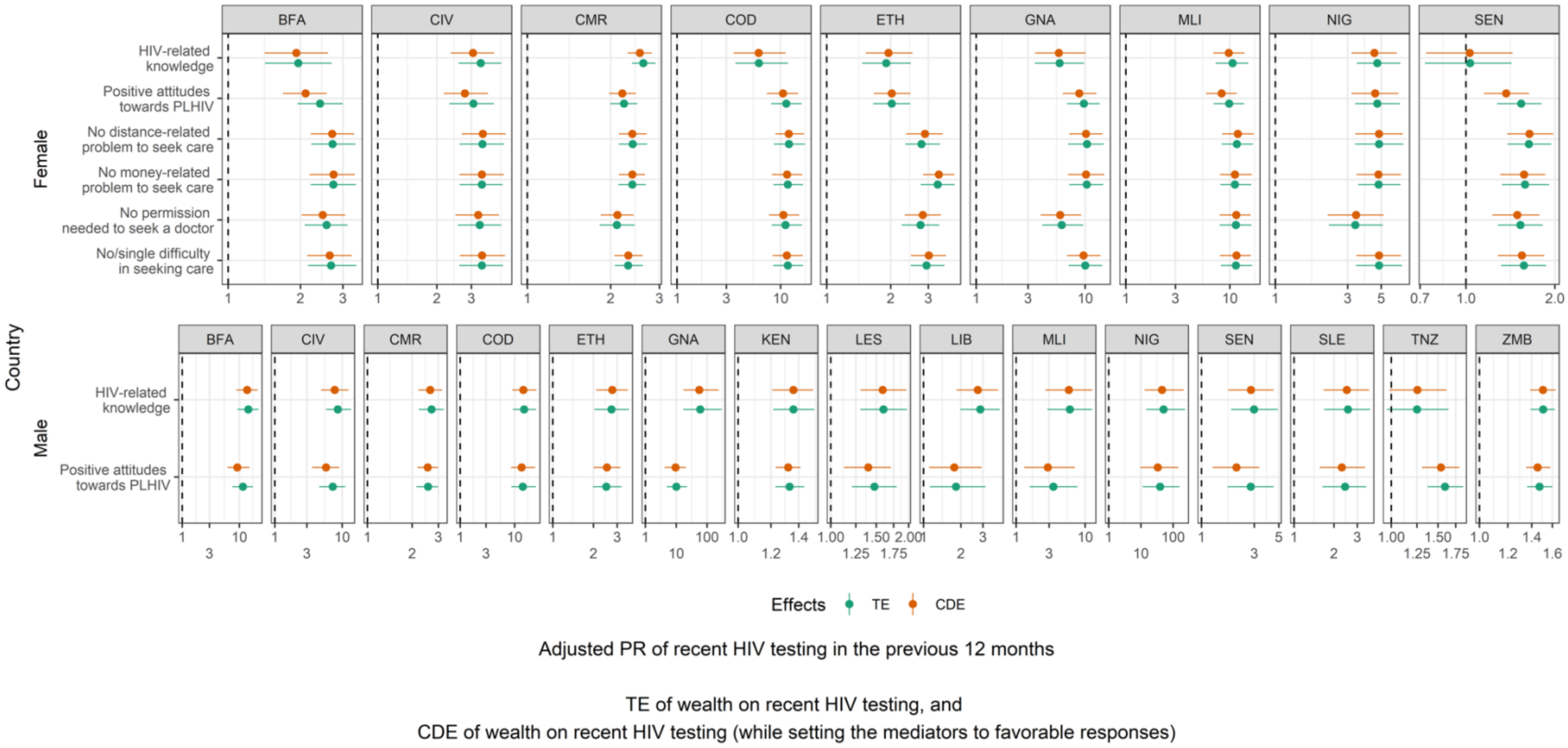

**Figure S3. Forest plot of the Total Effect and Controlled Direct Effect by mediator and gender.** Refer to Table S2 for full country names.

### Table S5. Proportion mediated by individual and joint mediators, stratified by gender, in 18 sub-Saharan African countries.

| Country † | Gender | HIV-related knowledge | Positive attitudes toward PLHIV | Sum | Mean | Joint demand-related mediator | No distance-related problem to seek care | No money-related problem to seek care | No permission needed to seek a doctor | Sum | Mean | Joint supply-related mediator |
|---|---|---|---|---|---|---|---|---|---|---|---|---|
| **BFA** | **Female** | 4.0% | 22.0% | 26.0% | 13.0% | 13.0% | 1.0% | 6.0% | 0.0% | 6.0% | 2.0% | 1.0% |
| | **Male** | 5.0% | 22.0% | 27.0% | 14.0% | 12.0% | - | - | - | - | - | - |
| **CIV** | **Female** | 12.0% | 14.0% | 26.0% | 13.0% | 10.0% | -1.0% | 3.0% | -0.0% | 2.0% | 1.0% | 0.0% |
| | **Male** | 12.0% | 24.0% | 36.0% | 18.0% | 20.% | - | - | - | - | - | - |
| **CMR** | **Female** | 5.0% | 3.0% | 7.0% | 4.0% | 3.0% | 1.0% | -1.0% | 0.0% | 0.0% | 0.0% | 0.0% |
| | **Male** | 3.0% | 1.0% | 4.0% | 2.0% | 2.0% | - | - | - | - | - | - |
| **COD** | **Female** | 0.0% | 8.0% | 8.0% | 4.0% | 4.0% | 0.0% | 5.0% | 2.0% | 7.0% | 2.0% | 1.0% |
| | **Male** | 3.0% | 5.0% | 8.0% | 4.0% | 5.0% | - | - | - | - | - | - |
| **ETH** | **Female** | -5.0% | 0.0% | -5.0% | -3.0% | -3.0% | -6.0% | -4.0% | -2.0% | -12.0% | -4.0% | -2.0% |
| | **Male** | -2.0% | -2.0% | -4.0% | -2.0% | -2.0% | - | - | - | - | - | - |
| **GNA** | **Female** | 2.0% | 10.0% | 12.0% | 6.0% | 6.0% | 2.0% | 4.0% | 1.0% | 8.0% | 3.0% | 2.0% |
| | **Male** | 8.0% | 6.0% | 13.0% | 7.0% | 6.0% | - | - | - | - | - | - |
| **KEN** | **Female** | 6.0% | 3.0% | 9.0% | 4.0% | 4.0% | -8.0% | -6.0% | -1.0% | -16.0% | -5.0% | -1.0% |
| | **Male** | 0.0% | 3.0% | 3.0% | 2.0% | 1.0% | - | - | - | - | - | - |
| **LES** | **Female** | -5.0% | -6.0% | -12.0% | -6.0 | -5.0% | 6.0% | -1.0% | 0.0% | 5.0% | 2.0% | -1.0% |
| | **Male** | 2.0% | 16.0% | 18.0% | 9.0% | 1.0% | - | - | - | - | - | - |
| **LIB** | **Female** | 2.0% | 32.0% | 34.0% | 17.0% | 32.0% | 14.0% | 4.0% | 1.0% | 19.0% | 6.0% | 5.0% |
| | **Male** | 7.0% | 7.0% | 15.0% | 7.0% | 7.0% | - | - | - | - | - | - |

† Refer to Table S2 for full country names.

### Table S5 (continued). Proportion mediated by individual and joint mediators, stratified by gender, in 18 sub-Saharan African countries.

| Country † | Gender | HIV-related knowledge | Positive attitudes toward PLHIV | Sum | Mean | Joint demand-related mediator | No distance-related problem to seek care | No money-related problem to seek care | No permission needed to seek a doctor | Sum | Mean | Joint supply-related mediator |
|---|---|---|---|---|---|---|---|---|---|---|---|---|
| **MLI** | **Female** | 9.0% | 17.0% | 26.0% | 13.0% | 14.0% | -3.0% | -1.0% | 0.0% | -4.0% | -1.0% | -1.0% |
| | **Male** | 4.0% | 25.0% | 29.0% | 15.0% | 17.0% | - | - | - | - | - | - |
| **MWI** | **Female** | 3.0% | 2.0% | 5.0% | 2.0% | 2.0% | 1.0% | 3.0% | 1.0% | 5.0% | 2.0% | 1.0% |
| | **Male** | 3.0% | 7.0% | 10.0% | 5.0% | 4.0% | - | - | - | - | - | - |
| **NIG** | **Female** | 5.0% | 4.0% | 10.0% | 5.0% | 7.0% | 0.0% | -2.0% | 0.0% | -2.0% | -1.0% | 0.0% |
| | **Male** | 10.0% | 15.0% | 26.0% | 13.0% | 16.0% | - | - | - | - | - | - |
| **RWA** | **Female** | 1.0% | 19.0% | 20.0% | 10.0% | 6.0% | -1.0% | 48.0% | 2.0% | 48.0% | 16.0% | 14.0% |
| | **Male** | -2.0% | -11.0% | -13.0 | -7.0% | -5.0% | - | - | - | - | - | - |
| **SEN** | **Female** | 7.0% | 31.0% | 39.0% | 19.0 | 26.0% | -1.0% | 7.0% | 2.0% | 8.0 | 3.0% | 3.0% |
| | **Male** | 9.0% | 40.0% | 49.0% | 25.0% | 28.0% | - | - | - | - | - | - |
| **SLE** | **Female** | 13.0% | 10.0% | 23.0% | 12.0% | 13.0% | 6.0% | 14.0% | 1.0% | 21.0% | 7.0% | 4.0% |
| | **Male** | 3.0% | 9.0% | 12.0% | 6.0% | 7.0% | - | - | - | - | - | - |
| **TNZ** | **Female** | 9.0% | 17.0% | 26.0% | 13.0% | 17.0% | - | - | - | - | - | - |
| | **Male** | -1.0% | 9.00% | 8.00% | 4.0% | 1.0% | - | - | -- | - | - | - |
| **ZBW** | **Female** | 9.0% | 6.0% | 15.0% | 7.0% | 7.0% | 5.0% | 0.2% | 0.1% | 0.3% | 0.1% | 0.1% |
| | **Male** | 6.0% | 4.0% | 10.0% | 5.0% | 5.0% | - | - | - | - | - | - |
| **ZMB** | **Female** | 5.0% | -10.0% | -6.0% | -3.0% | -3.0% | -178.0% | 1.0% | 0.0% | -177.0% | -59.0% | -1.0% |
| | **Male** | 0.0% | 4.0% | 4.0% | 2.0% | 2.0% | - | - | - | - | - | - |

† Refer to Table S2 for full country names.
